# Perceived Stress and Psychological (Dis)Stress among Indian Endodontists During COVID19 Pandemic Lock down

**DOI:** 10.1101/2020.05.06.20092601

**Authors:** RN Kumar Anil, SC Karumaran, Deepthi Kattula, Rooban Thavarajah, AM Anusa

## Abstract

**Background:** The novel 2019 coronavirus(COVID-19) spreads by respiratory and aerosols. COVID19 driven pandemic causes panic, fear and stress among all strata of society. Like all other medical professions, dentists, particularly endodontists, who are highly exposed to aerosols would be exposed to stress. The aim of this study was to assess the (dis)stress among Indian endodontists and the factors that could influence the (dis)stress.

**Methods:** From 8^th^ April to 16^th^ April 2020, we conducted an online survey in closed endodontic social media using snowball sampling technique, collecting basic demographic data, practice setting and relevant data. Psychological stress and perceived distress were collected through COVID-19 Peri-traumatic Distress Index (CPDI) and Perceived stress scale (PSS). Multinomial regression analysis was performed to estimate relative risk rate and P≤0.05 was considered significant.

**Results:** This study had 586 Indian endodontists completing this survey across India. Of these, 311 (53.07%) were males, 325(55%) in the age group of 25-35 years, 64%in urban areas, 13.14% in solo-practice and a fourth of them were residents. Female endodontists had high perceived stress (RRR=2.46,P=0.01) as compared to males, as measured by PSS. Younger endodontists<25 years(RRR=9.75;P=0.002) and 25-35years (RRR=4.60;P=0.004) as compared with >45 years age-group had more distress. Exclusive consultants had RRR= 2.90, P=0.02, for mild-to-moderate distress as compared to normal. Factors driving this phenomenon are considered.

**Conclusions:** During the lock down due to COVID-19, 1-in-2 Indian endodontists had distress, as measured by CPDI and 4-in-5 of them had perceived stress, as indicated by PSS. Our model identified certain factors driving the (dis)stress, which would help policy framers to initiate appropriate response.

## BACKGROUND

The current pandemic, attributed to a novel severe acute respiratory syndrome coronavirus 2 (SARS CoV2) better known as COVID-19, has affected dental practice, like all other medical services. This virus strain ears a striking resemblance to the 21^st^ century viruses like SARS-CoV and Middle East respiratory syndrome coronavirus(MERS-CoV). COVID19 is reported to be a zoonotic virus, with bats being the possible origin and pangolins as probable intermediate host. The first transmission probably occurred in an exotic meat rich wet market in Wuhan, China. Following this animal-to-human transmission, the COVID19 has been spread by human-to-human contact. The interpersonal contact occurs mainly via respiratory droplets/aerosols, saliva and or contact. The role of fecal-oral transmission, through fomites or vertical transmission are yet to be confirmed. Both symptomatic and asymptomatic patients spread the virus, but the latter, may act as super-spreaders, as they do not have symptoms and remain active in the community. The incubation period of COVID-19 is reported to be about 5 to 6 days, with up to 14 days have been reported and this is widely used as cut off days for medical observation and quarantine.(Meng et al., 2020)

In India, the first confirmed case of COVID-19 was reported on January 30, 2020.(Gupta et al., 2020) As a containment initiative, the nation was placed on a total lock down since early hours of 25^th^ March 2020.(Pulla, 2020) With the start of phase-1 lockdown, all diagnosed cases of COVID19 underwent contact tracing and potential contacts, including Health Care Workers (HCW) were quarantined or isolated. As of the writing of this article, the nation continues to be under lockdown as phase-2, till 3^rd^ May 2020 and the total number of cases on 22^nd^ April 2020 morning, was 19984 COVID19 positive cases including 640 deaths (Source: www.mohfw.gov.in)

Oral health care involves use of aerosol producing devices as well as working in close proximity with the oral mucosa and secretions. The risk of cross infection could be high between dental practitioners and COVID19 patients. (Dave et al., 2020; Martelli-Júnior et al., 2020) Following the universal norms, societal and government advisory, most of the dental practices and hospitals in India, have closed operation since 25^th^ March. Only emergency and urgent cases were taken up. Even in such instances, strict and effective infection control protocols were to be followed.(Amber et al., 2020; Dave et al., 2020; Farooq and Ali, 2020) In spite of such universal precautions, when a COVID19 patient or a carrier is treated, the risk of aerosol induced spread of virus appears to higher, creating numerous hot-sites of virus deposition in the operatory. (Guo et al., 2020; Veena et al., 2015) Among dentists, there is a constant, looming anxiety of encountering a COVID19 infected patient especially when there is lack or limited of access to personal protective equipment, no proper standard protocol for management and possibility of incurring financial implications in the future due to decreased clinical operation hours.(Farooq and Ali, 2020)

Endodontists, who are dental specialists (trained primarily to save the decayed tooth, often through root canal treatment) are at greater risk to encounter the cross-infection because of employment of high-speed rotary instruments generating large volume of aerosols and splatter of saliva, during treatment, increasing the probability of nosocomial spread of COVID19. This could endanger endodontists, their patients and the community that they serve.(Amber et al., 2020; Peng et al., 2020)

Widespread of disease, in short time, creates stress and has a psychological impact. In the present COVID 19 crisis, a Chinese general population survey documented more than a third of the respondents manifested psychological stress with 5% experiencing severe distress.(Qiu et al., 2020) Among Indians, in a small general public survey, 80 % of the respondents were preoccupied with the thoughts of COVID-19, while sleep difficulties (12.5%), paranoid about acquiring COVID-19 infection (37.8%) and distress (36.4%) were also reported. There was a perceived mental healthcare need in at least 80 % of participants.(Roy et al., 2020)

A study among Chinese HCW exposed to COVID-19 patients reported distress(75%), depression(50%), anxiety(45%), and insomnia(34%).(Lai et al., 2020) During the SARS COV pandemic, HCW feared contagion and infection of their family, friends, colleagues, were reluctant to work and even contemplated resignation.(Bai et al., 2004) They experienced great deal of stress, anxiety and depression due to lurking uncertainty.(McAlonan et al., 2007)^]^

Till date, the psychological stress perception due to COVID19 has not been assessed among Indian endodontists though they fall under high-risk group for this infection. This manuscript intends to address this lacunae, from this part of the world. The present study evaluated the patterns of perceived psychological (dis)stress among the Indian endodontists and understand the factors associated with it.

## MATERIALS AND METHODS

A pan-Indian online survey for assessing the psychological stress among Indian endodontists was organized during 8^th^-16^th^ April, 2020. This coincided with the last days of phase1 of Indian lock down and initial days of the extension of lockdown. The exemption for this survey was sought from primary author’s institutional review committee as this was an anonymous survey with no personal identification collected out and carried out during a humanitarian emergencies and disaster, as per Indian Council of Medical Research guidelines.

This self-reportable form in English was designed using simple Google forms and the link shared among Indian endodontists through several endodontic specific, closed social media forums, using snowball-sampling technique. Anonymity was ensured and no personal identification, such as IP address, emailIDs or details of COVID19 exposure were collected. The basic demographics of gender, age group in years (>25, 25 to 35, SB-45, 45 and above), dental practice setting (solo practice-stand-alone practice engaging in endodontics and all dental treatment; private solo practice with other endodontic consultations and or part of a group practice (PSPC/GP); Consultations - consulting for endodontics exclusively at more than 1 clinic; Teaching and Practice - engaged in academics as well as clinical endodontic practice privately; Trainee under supervision - Residents or postgraduates of endodontics at a recognized dental institution, leading to master’s degree in endodontics) and experience after MDS (Trainee/PG student in endodontics, 0 to 5 years, 6 to 10 years and above 10 years) in years were collected. This survey used 2 previously validated questionnaires - COVID-19 Peritraumatic Distress Index (CPDI) and Perceived stress scale (PSS).(Cohen et al., 1983; Qiu et al., 2020)

The CPDI scale measured the frequency of anxiety, depression, specific phobias, cognitive change, avoidance and compulsive behavior, physical symptoms and loss of social functioning in the past week, quantifying on a scale of 0 to 100. A CPDI score of ≤27 had low or no distress, 28 to 51 indicates mild to moderate distress and score ≥52 indicates severe distress.(Qiu et al., 2020) The PSS inquired about thoughts and feelings, quantified as a scale of 0 to 40 with a score between 0-13 as low perceived stress(PS), 14-2B as moderate PS and 27-40 was considered to have high PS.(Cohen et al., 1983)

The data thus captured were entered and analyzed using Stata software, version 14.1 for Windows (Stata Corp, College Station, Texas). Descriptive statistics mean with standard deviation (SD), median for continuous variables and proportions with 95% confidence interval (CI) for categorical variables was calculated. Association of demographic variables with PS and distress levels obtained by PSS and CPDI scales is assessed by Pearson χ^2^ test. Magnitude of association of demographic variable with levels of PS/distress was analyzed using multinomial logistic regression and presented as relative risk ratios (RRR) with 95%CI. Hosmer-Lemeshow goodness-of fit-test was used to assess the model fit. The correlation between the two scales was analyzed using Pearson correlation test. An attempt was also made to differentiate the (dis)stress levels between the phase-1 period (8^th^ April to 14^th^ April 10:00 AM) and after the phase-2 lock down announcement (after 14^th^ April 10:05 AM) using 2-sample T test. In all instances, P<0.05 were considered statistically significant.

## RESULT

This survey received a total of 586 valid responses from endodontists across India during the study period. Of them 311 (53.07%) were males and 325(55%) of the respondents belonged to the age group of 25-35 years. Predominantly the endodontists practiced in urban area(64%), engaging in solo practice(13.14%), private & group practice along with consultations (PSPC/GP-23.21%), only consultations(8.02%) and academics along with clinical practice(27.65%). Twenty seven percent of the respondents were endodontic postgraduate students. One fourth (2 6.79%) of the respondents had more than 10 years of post Master Degree experience. Among the participants, only 6.48% used rubber dam always in their regular practice.(Table-1)

**Table-1:**
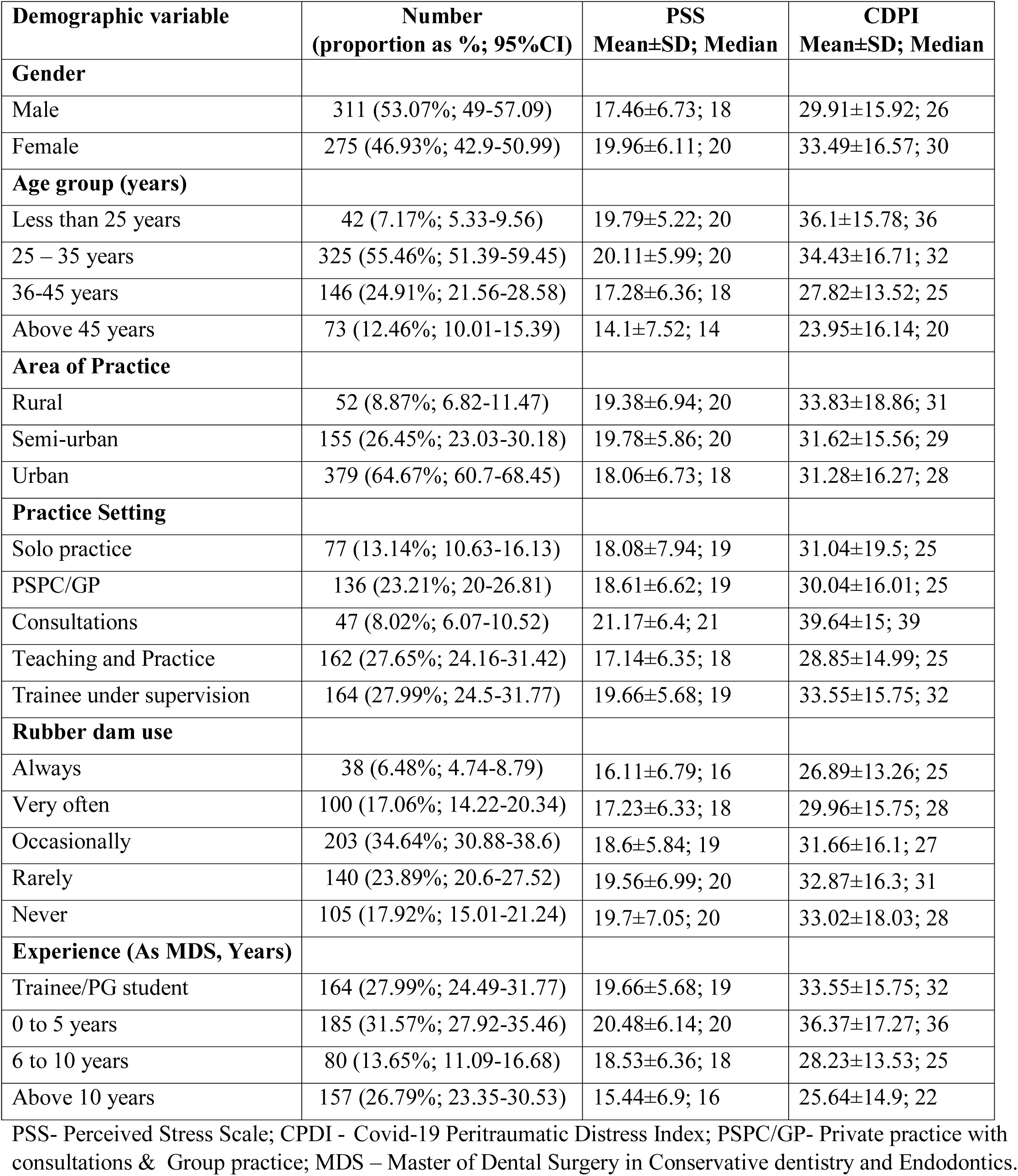
Demographics of Study population & Distribution of PSS, CPDI scores

| **Demographic variable** | **Number**<br>**(proportion as %; 95%CI)** | **PSS**<br>**Mean±SD; Median** | **CDPI**<br>**Mean±SD; Median** |
| --- | --- | --- | --- |
| **Gender** |  |  |  |
| Male | 311 (53.07%; 49-57.09) | 17.46±6.73; 18 | 29.91±15.92; 26 |
| Female | 275 (46.93%; 42.9-50.99) | 19.96±6.11; 20 | 33.49±16.57; 30 |
| **Age group (years)** |  |  |  |
| Less than 25 years | 42 (7.17%; 5.33-9.56) | 19.79±5.22; 20 | 36.1±15.78; 36 |
| 25 – 35 years | 325 (55.46%; 51.39-59.45) | 20.11±5.99; 20 | 34.43±16.71; 32 |
| 36-45 years | 146 (24.91%; 21.56-28.58) | 17.28±6.36; 18 | 27.82±13.52; 25 |
| Above 45 years | 73 (12.46%; 10.01-15.39) | 14.1±7.52; 14 | 23.95±16.14; 20 |
| **Area of Practice** |  |  |  |
| Rural | 52 (8.87%; 6.82-11.47) | 19.38±6.94; 20 | 33.83±18.86; 31 |
| Semi-urban | 155 (26.45%; 23.03-30.18) | 19.78±5.86; 20 | 31.62±15.56; 29 |
| Urban | 379 (64.67%; 60.7-68.45) | 18.06±6.73; 18 | 31.28±16.27; 28 |
| **Practice Setting** |  |  |  |
| Solo practice | 77 (13.14%; 10.63-16.13) | 18.08±7.94; 19 | 31.04±19.5; 25 |
| PSPC/GP | 136 (23.21%; 20-26.81) | 18.61±6.62; 19 | 30.04±16.01; 25 |
| Consultations | 47 (8.02%; 6.07-10.52) | 21.17±6.4; 21 | 39.64±15; 39 |
| Teaching and Practice | 162 (27.65%; 24.16-31.42) | 17.14±6.35; 18 | 28.85±14.99; 25 |
| Trainee under supervision | 164 (27.99%; 24.5-31.77) | 19.66±5.68; 19 | 33.55±15.75; 32 |
| **Rubber dam use** |  |  |  |
| Always | 38 (6.48%; 4.74-8.79) | 16.11±6.79; 16 | 26.89±13.26; 25 |
| Very often | 100 (17.06%; 14.22-20.34) | 17.23±6.33; 18 | 29.96±15.75; 28 |
| Occasionally | 203 (34.64%; 30.88-38.6) | 18.6±5.84; 19 | 31.66±16.1; 27 |
| Rarely | 140 (23.89%; 20.6-27.52) | 19.56±6.99; 20 | 32.87±16.3; 31 |
| Never | 105 (17.92%; 15.01-21.24) | 19.7±7.05; 20 | 33.02±18.03; 28 |
| **Experience (As MDS, Years)** |  |  |  |
| Trainee/PG student | 164 (27.99%; 24.49-31.77) | 19.66±5.68; 19 | 33.55±15.75; 32 |
| 0 to 5 years | 185 (31.57%; 27.92-35.46) | 20.48±6.14; 20 | 36.37±17.27; 36 |
| 6 to 10 years | 80 (13.65%; 11.09-16.68) | 18.53±6.36; 18 | 28.23±13.53; 25 |
| Above 10 years | 157 (26.79%; 23.35-30.53) | 15.44±6.9; 16 | 25.64±14.9; 22 |

PSS- Perceived Stress Scale; CPDI - Covid-19 Peritraumatic Distress Index; PSPC/GP- Private practice with consultations & Group practice; MDS – Master of Dental Surgery in Conservative dentistry and Endodontics.

The correlation coefficient (r) between PSS and CPDI was 0.627 with P<0.0001.The Cronbach’s ***α*** score for PSS was 0.8 and 0.921 for the CPDI. The mean (SD) score for PSS and CPDI was 17.46(6.73) and 29.91(15.92) respectively. PSS documented perceived moderate PS(score 14-26) among 406(69.28%) and high PS in 63(10.75%) respondents, whereas CPDI recorded 224(38.23%) and 80(13.65%) experienced mild-moderate and severe distress respectively (Figure-1). The distribution of mean (SD), median PSS and CPDI scores for demographic variables are provided in Table-1.

**Figure-1.**
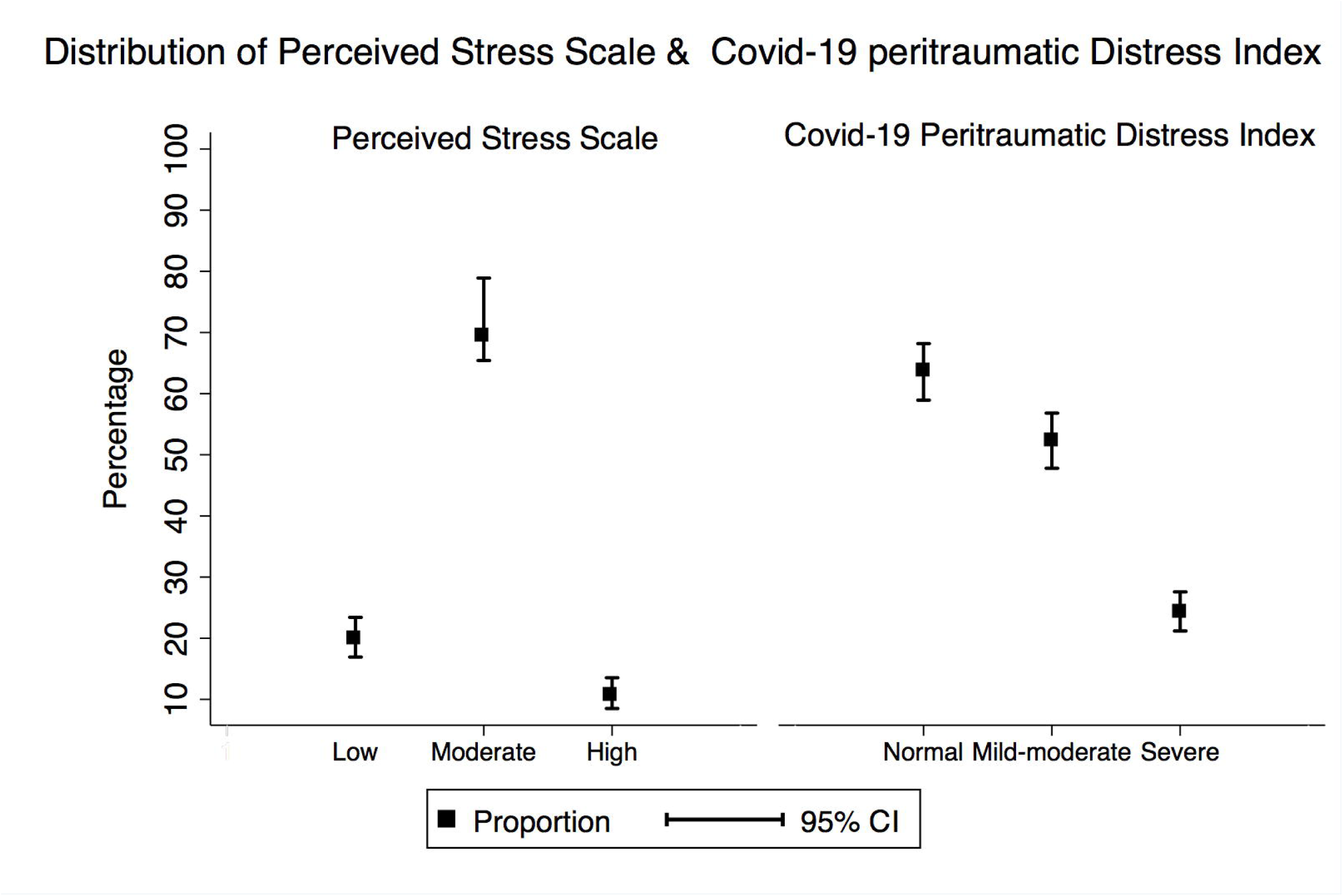

An initial analysis using contingency test based on the Pearson χ^2^ test was conducted to identify the association of stress/distress levels with demographic factors such as gender, age group, practice setting and post Master Degree experience. Almost all the demographic factors were significantly (P<0.05) associated with the different levels in PSS and CPDI scales(Table-2). As rubber dam usage was skewed among the respondents, it was not considered for further analysis as it could seed bias. As place of practice was not significant among CPDI, it was removed from further analysis.

**Table-2:** Association of demographics characteristics with various levels of PSS and CPDI scale

| Characteristic<br>s | Perceived Stress Scale<br>N(%) |  |  |  | Covid-19 Peritraumatic Distress Index<br>N(%) |  |  |  |
| --- | --- | --- | --- | --- | --- | --- | --- | --- |
|  | Low | Moderate | High | P value | Normal | Mild to<br>moderate | Severe | P value |
| **Gender** |  |  |  |  |  |  |  |  |
| Male | 78(66.7) | 210 (51.72) | 23(36.51) | <0.0001 | 167<br>(59.22) | 109 (48.66) | 35 (43.75) | 0.01 |
| Female | 39(33.3) | 196 (48.28) | 40 63.49) |  | 115<br>(40.78) | 115 (51.34) | 45 (56.25) |  |
| **Age group** |  |  |  |  |  |  |  |  |
| <25 years | 6 (5.13) | 30(7.39) | 6(9.52) | <0.0001 | 13(4.61) | 21(9.38) | 8(10.0) | <0.0001 |
| 25-35 years | 39(33.33) | 243(59.85) | 43(68.25) |  | 132<br>(46.81) | 134(59.82) | 59(73.75) |  |
| 36-45 years | 37(31.62) | 99(24.38) | 10(15.87) |  | 83(29.43) | 55(24.55) | 8(10.0) |  |
| >45 years | 35(29.91) | 34(8.37) | 4(6.35) |  | 54(19.15) | 14(6.25) | 5(6.25) |  |
| **Area of Practice** |  |  |  |  |  |  |  |  |
| Rural | 10(8.55) | 36(8.87) | 6(9.52) | 0.04 | 22(7.80) | 20(8.93) | 10(12.50) | 0.68 |
| Semi-urban | 18(15.38) | 118(29.06) | 19(30.16) |  | 73(25.89) | 59(26.34) | 23(28.75) |  |
| Urban | 89(76.07) | 252(62.07) | 38(60.32) |  | 187<br>(66.31) | 145(64.73) | 47(58.75) |  |
| **Practice setting** |  |  |  |  |  |  |  |  |
| Solo practice | 23(19.66) | 46(11.33) | 8(12.70) | <0.0001 | 41(14.54) | 22(9.82) | 14(17.50) | <0.0001 |
| PSPC/GP | 26(22.22) | 95(23.40) | 15(23.81) |  | 76(26.95) | 44(19.64) | 16(20.0) |  |
| Consultations | 4(3.42) | 33(8.13) | 10(15.87) |  | 11(3.90) | 21(9.38) | 15(18.75) |  |
| Teaching &<br>practice | 45(38.46) | 107(26.35) | 10(15.87) |  | 91(32.27) | 56(25.0) | 15(18.75) |  |
| Training under<br>supervision | 19(16.24) | 125(30.79) | 20(31.75) |  | 63(22.34) | 81(36.16) | 20(25.0) |  |
| **Experience (as MDS, Years)** |  |  |  |  |  |  |  |  |
| Trainee/PG<br>student | 19(16.24) | 125(30.79) | 20(31.75) | <0.0001 | 63(22.34) | 81(36.16) | 20(25.0) | <0.0001 |
| 0 to 5 years | 21(17.95) | 135(33.25) | 29(46.03) |  | 68(24.11) | 71(31.70) | 46(57.50) |  |
| 6 to 10 years | 16(13.68) | 58(14.29) | 6(9.52) |  | 50(17.73) | 24(10.71) | 6(7.50) |  |
| Above 10 years | 61(52.14) | 88(21.67) | 8(12.70) |  | 101<br>(35.82) | 48(21.43) | 8(10.0) |  |

PSS- Perceived Stress Scale; CPDI - Covid-19 Peritraumatic Distress Index; PSPC/GP- Private practice with consultations & Group practice; MDS – Master of Dental Surgery in Conservative dentistry and Endodontics.

In the univariate analysis, demographic factors associated with PSS scale (comparison of high vs. low PS) showed females(RRR=3.47;P<0.001), specialists employed through only consultations (RRR=7.18;P=0.006), postgraduate students under supervised training (RRR=3.02;P=0.03), individuals in the age group of <25 years (RRR=8.75;P= 0.006), 25-35 years (RRR=9.64;P< 0.001), specialists with <5 years of work experience (RRR=10.52;P<0.0001) and postgraduate students (RRR=8.02;P<0.0001) were significantly associated with increased risk for high PS. Similar trend of association with demographic factors was observed when moderate PS was compared with low PS. (Table-3) Multivariate analysis was carried out assessing the factors influencing the PSS scale, for the high PS, as compared to low PS. The model fit test(χ2=45.41, P=0.41), suggested that the model showed adequately fit for the covariates used. Female endodontists (RRR=2.46,P=0.01) were significantly associated with an increased risk of high PS as compared to males. Although specialist employed through only consultations was associated with risk of high PS (RRR=3.59, P=0.08), it was statistically at borderline significance. Younger age group (25-35years) individuals were observed to experience increased high PS in comparison with older age group (>45 years) and findings were statistically significant (Table-4).

**Table-3.** Results of UNIVARIATE ANALYSIS with Demographics vs PSS and CPDI scales

|  | PSS (RRR (95%CI); P-Value |  | CPDI (RRR (95%CI); P-Value |  |
| --- | --- | --- | --- | --- |
|  | Moderate Vs Low stress | High Vs Low Stress | Mild-moderate vs Normal Stress | Severe vs Normal Stress |
| **Gender** |  |  |  |  |
| Females | 1.86 (1.21 - 2.87);<br>0.005 | 3.47 (1.83 - 6.6);<br><0.0001 | 1.53 (1.07 - 2.18);<br>0.02 | 1.86 (1.13 - 3.08);<br>0.01 |
| Males |  |  |  |  |
| **Age Group** |  |  |  |  |
| <25 years | 5.15(1.90 - 13.92);<br>0.001 | 8.75(1.88 - 40.53);<br>0.006 | 6.23(2.51 -15.44);<br><0.0001 | 6.64(1.86 -23.68);<br>0.003 |
| 25 to 35 years | 6.41(3.58 -<br>11.46);<0.0001 | 9.64 (3.14 -<br>29.61); <0.0001 | 3.91(2.07 - 7.39);<br><0.0001 | 4.82(1.83 -12.68);<br>0.001 |
| 36-45 years | 2.75(1.50 -5.04);<br>0.001 | 2.36 (0.67 - 8.23);<br>0.17 | 2.55 (1.29 -5.04) ;<br>0.007 | 1.04(0.32-3.34);<br>0.94 |
| Above 45 years |  |  |  |  |
| **Area of Practice** |  |  |  |  |
| Semi-urban | 1.82 (0.77 - 4.29);<br>0.17 | 1.75 (0.53 - 5.84);<br>0.35 | 0.88 (0.44 - 1.78);<br>0.74 | 0.69 (0.28 - 1.67);<br>0.41 |
| Urban | 0.78 (0.37 -1.65);<br>0.52 | 0.71 (0.24 -2.09);<br>0.53 | 0.85 (0.44 - 1.62);<br>0.62 | 0.55 (0.24 -1.24);<br>0.15 |
| Rural |  |  |  |  |
| **Practice Setting** |  |  |  |  |
| Solo practice |  |  |  |  |
| PSPC/GP | 1.82 (0.94 - 3.54);<br>0.07 | 1.65 (0.59 - 4.62);<br>0.33 | 1.07 (0.57- 2.04);<br>0.81 | 0.61 (0.27- 1.38);<br>0.24 |
| Consultations | 4.12 (1.3 -13.05);<br>0.01 | 7.18 (1.75 - 29.4);<br>0.006 | 3.55 (1.45- 8.7);<br>0.005 | 3.99 (1.48 -10.71);<br>0.006 |
| Teaching & practice | 1.18 (0.64 -2.18);<br>0.57 | 0.63 (0.22 -1.83);<br>0.4 | 1.14 (0.61-2.12);<br>0.66 | 0.48 (0.21 -1.09);<br>0.08 |
| Training under supervision | 3.28 (1.64 - 6.59);<br>0.001 | 3.02 (1.09 - 8.39);<br>0.03 | 2.39 (1.29 - 4.42);<br>0.005 | 0.92 (0.42 - 2.04);<br>0.85 |
| Solo Practice |  |  |  |  |
| **Experience (as MDS in years)** |  |  |  |  |
| Trainee/PG student | 4.56(2.54 - 8.16);<br><0.0001 | 8.02(3.04 - 21.13);<br><0.0001 | 2.70(1.68-4.35);<br><0.0001 | 4.01(1.66- 9.64);<br>0.002 |
| 0 to 5 years | 4.5(2.53 -7.83);<br><0.0001 | 10.52(4.16 -<br>26.59); p<0.0001 | 2.19(1.36 -3.54);<br><0.001 | 8.54(3.79 - 19.22);<br><0.0001 |
| 6 to 10 years | 2.51(1.32 - 4.77);<br>0.005 | 2.85(0.86 - 9.42);<br>0.08 | 1.01(0.55 - 1.83);<br>0.97 | 1.51(0.49 - 4.60);<br>0.46 |
| Above 10 years |  |  |  |  |

PSS- Perceived Stress Scale; CPDI - Covid-19 Peritraumatic Distress Index; RRR- Relative Risk Rate; PSPC/GP- Private practice with consultations & Group practice; MDS – Master of Dental Surgery in Conservative dentistry and Endodontics.

**Table-4:**
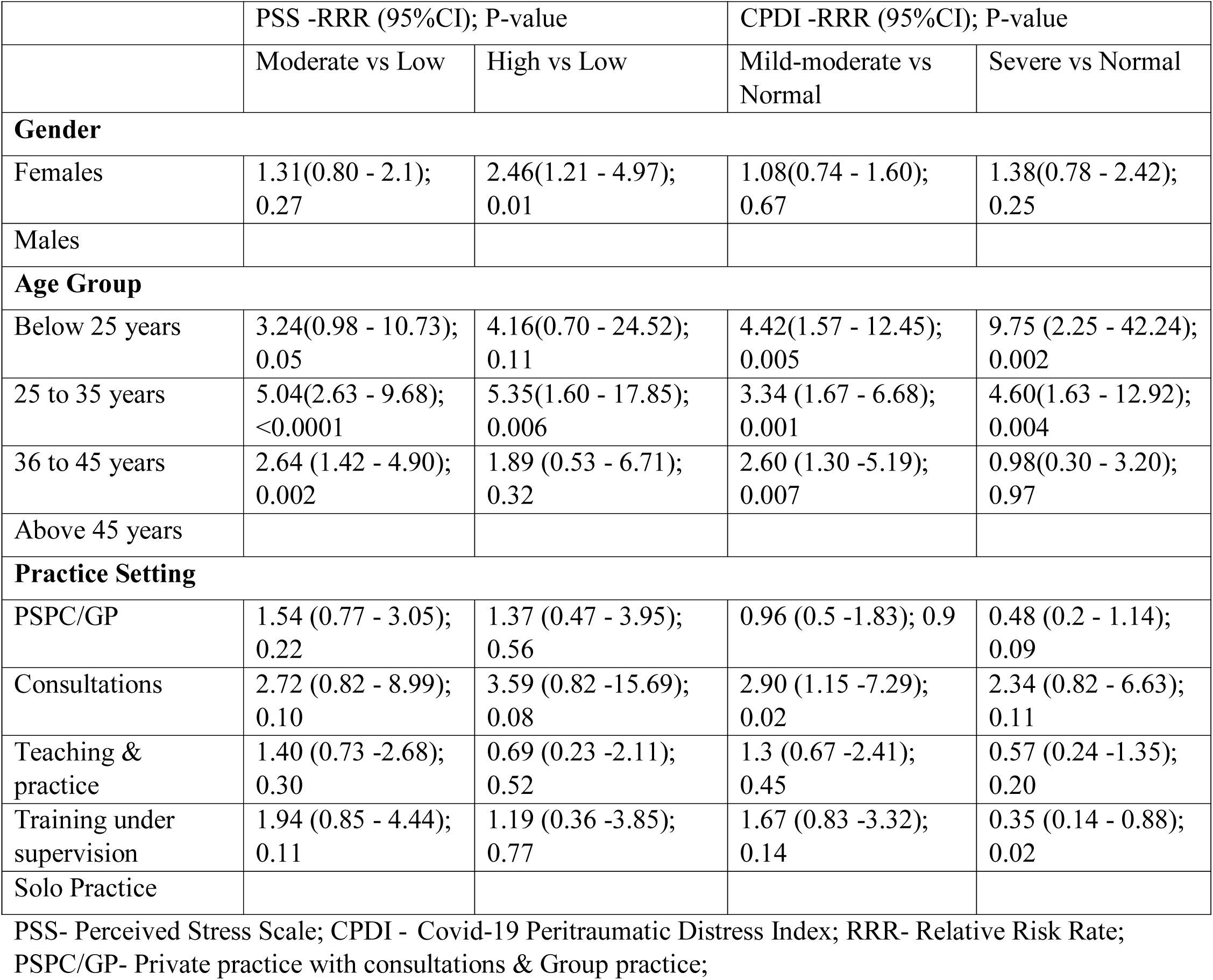
Multivariate Demographics vs PSS and CPDI scales

|  | PSS -RRR (95%CI); P-value |  | CPDI -RRR (95%CI); P-value |  |
| --- | --- | --- | --- | --- |
|  | Moderate vs Low | High vs Low | Mild-moderate vs Normal | Severe vs Normal |
| **Gender** |  |  |  |  |
| Females | 1.31(0.80 - 2.1);<br>0.27 | 2.46(1.21 - 4.97);<br>0.01 | 1.08(0.74 - 1.60);<br>0.67 | 1.38(0.78 - 2.42);<br>0.25 |
| Males |  |  |  |  |
| **Age Group** |  |  |  |  |
| Below 25 years | 3.24(0.98 - 10.73);<br>0.05 | 4.16(0.70 - 24.52);<br>0.11 | 4.42(1.57 - 12.45);<br>0.005 | 9.75 (2.25 - 42.24);<br>0.002 |
| 25 to 35 years | 5.04(2.63 - 9.68);<br><0.0001 | 5.35(1.60 - 17.85);<br>0.006 | 3.34 (1.67 - 6.68);<br>0.001 | 4.60(1.63 - 12.92);<br>0.004 |
| 36 to 45 years | 2.64 (1.42 - 4.90);<br>0.002 | 1.89 (0.53 - 6.71);<br>0.32 | 2.60 (1.30 -5.19);<br>0.007 | 0.98(0.30 - 3.20);<br>0.97 |
| Above 45 years |  |  |  |  |
| **Practice Setting** |  |  |  |  |
| PSPC/GP | 1.54 (0.77 - 3.05);<br>0.22 | 1.37 (0.47 - 3.95);<br>0.56 | 0.96 (0.5 -1.83); 0.9 | 0.48 (0.2 - 1.14);<br>0.09 |
| Consultations | 2.72 (0.82 - 8.99);<br>0.10 | 3.59 (0.82 -15.69);<br>0.08 | 2.90 (1.15 -7.29);<br>0.02 | 2.34 (0.82 - 6.63);<br>0.11 |
| Teaching & practice | 1.40 (0.73 -2.68);<br>0.30 | 0.69 (0.23 -2.11);<br>0.52 | 1.3 (0.67 -2.41);<br>0.45 | 0.57 (0.24 -1.35);<br>0.20 |
| Training under supervision | 1.94 (0.85 - 4.44);<br>0.11 | 1.19 (0.36 -3.85);<br>0.77 | 1.67 (0.83 -3.32);<br>0.14 | 0.35 (0.14 - 0.88);<br>0.02 |
| Solo Practice |  |  |  |  |

PSS- Perceived Stress Scale; CPDI - Covid-19 Peritraumatic Distress Index; RRR- Relative Risk Rate; PSPC/GP- Private practice with consultations & Group practice;

Univariate analysis of demographic factors associated with CPDI scale (comparison of severe vs. normal distress) documented females (RRR=1.86;P=0.01), individuals in the age group of <25 years (RRR=6.64; P=0.003), 25-35 years (RRR=4.82; P=0.001), endodontists consulting at clinics only (RRR=3.99;P= 0.006), specialist with less than 5 years of post Master Degree experience (RRR=8.54;P<0.0001) and postgraduate students (RRR=4.01; P=0.002) experienced significantly severe distress level. Comparison of mild to moderate with normal distress estimated similar magnitude of associations (Table-3). A model built for multivariate analysis showed adequate fit (*χ*^2^=59.46, P=0.06) for the covariates used. In the multivariate analysis, younger age, particularly among the age group of <25 (RRR=9.75; P = 0.002), 25-35 years (RRR=4.60; P= 0.004) in comparison with >45 years age and postgraduate students under supervised training (RRR=0.35, P= 0.02) in comparison with solo practitioners had significantly higher risk of severe distress. Specialist employed through only consultations was associated with higher risk for both mild to moderate (RRR= 2.90, P=0.02) with statistical significance while the severe distress (RRR= 2.34, P=0.11), was not statistically significant (Table-4).

The mean PSS score in phase-1 period (n=515) was 18.28±6.45(17.72-18.84) while the same after announcement of phase-2 lockdown (n=71) was 21.18±6.86(19.56-22.81) with the P=0.0005. For the CDPI, the scores were 30.96±15.85(29.59-32.33) and 36.18±18.87(31.72-40.65) respectively with P=0.0113.

## DISCUSSION

The social or physical distancing and other COVID19 restrictions have caused significant disruption globally. This disruptions extends from individuals to families, communities, and whole countries. This disruption also has altered previous familiar concepts and added complexities in all walks of life.(Usher et al., 2020) Mental health issues among HCW in COVID19 situation has been documented.(Greenberg et al., 2020; Kang et al., 2020) This altered mental health status stems from several aspects of disturbance from routine. This includes social isolation, interpersonal distancing, heightened need for infection control procedures domestics and at dental operatory, fear of contagion, public perception of COVID19 stigma, concerns for self/family wellbeing, potential of acing emotionally charged clinical environments, possible unrealistic public expectations, prevalent cynicism, possibility of patient safety incidents, procedural errors, decreased access to protection gears, lower quality of para-service provision, along with core issues of financial insecurity and potential loss of income.(Gavin et al., 2020) Economic impact of COVID19 pandemic is at a personal, community, national and global level. This burden cannot be underestimated, as it could potentially influence on all other spheres of life.(Mckee and Stuckler, 2020) Dentists and Endodontists, as an integral part of the society, are not immune from all these impacts. There is dearth of evidence of perceived stress (PS) and distress among dentists, particularly among endodontists, (irrespective of COVID19 exposure) who are at the highest risk of cross infection.(Coulthard, 2020) For this very purpose, we used two scales, a time tested PSS scale and new, COVID19 specific CPDI scale were used. Both have been used and published in peer reviewed literature and been accepted, with the former being used for more than three decades.(Cohen et al., 1983; Qiu et al., 2020) As there was a statistically significant correlation, it could be safely inferred that the COVID19 scale is reflective of the self-reported psychological distress in COVID19 situation. However, as the PSS scores have not been used previously in COVID19 related literature, rendering it impossible to be compared with pertinent literature in this manuscript.

The mean score of CPDI for this exclusive Indian endodontic cohort was 29.91±15.92 which is greater than the Chinese general population 23.65±15.45 but lower than that of Iranian population (34.54±14.92).(Afshar Jahanshahi et al., 2020; Qiu et al., 2020) The psychological distress among Indian endodontists, during the time of COVID19 by CPDI was 51.88% (38.23% mild to moderate and 13.65%-severe distress) while estimates of distress from Chinese population (by CPDI) was 35% (29.29% mild-to-moderate; 5.14% severe) and Iranians was 61 %(47.0% mild-to-moderate; 14.1% severe). The difference could be multifold – primarily that the present study had a homogenous cohort of endodontists whereas the Chinese and Iranian cohorts comprised of general public. The other reason is that Chinese and Iranian had a significant number of population infected with COVID19, unlike India.(Afshar Jahanshahi et al., 2020; Gupta et al., 2020, Pulla, 2020) The biological, anthropological, immunological, socio-demographic and cultural variations could also be accountable. Like the Chinese study, Indian Women endodontists had higher extent of stress while age group is not in agreement with Chinese study. (Qiu et al., 2020) Like the Iranian finding, the predictors of distress in India vary from those in China and Iran.(Afshar Jahanshahi et al., 2020; Qiu et al., 2020) When the PSS was employed, 80%(69.28%-moderate; 10.75%-severe) of Indian endodontists suffered from stress during COVID19 situation. This is 235% higher than that of the Indian general public during the same situation.(Roy et al., 2020) The PS proportions are similar to Chinese HCW who were exposed to COVID19 patients.(Lai et al., 2020)

Rubber dam usage among Indian endodontics is skewed, with only 6.48% of the cohort regularly using it. Also, there are conflicting evidences about the rubber dam’s efficacy in alteration microbial load in aerosols as compared to non-rubber dam use.(Al-Amad et al., 2017; Samaranayake et al., 1989) Hence, rubber dam usage characteristic was not used in further analysis.

The association of gender, age group, area of practice, practice setting and experience were statistically significant between the levels of (dis)stress using PSS and CPDI. (Table-2) As there is no pertinent literature, the present results cannot be supported or refuted.

In the univariate analysis, significance with gender reflects the fact that women, particularly female doctors are vulnerable to distress, which is similar to Chinese cohort.(Bhatia, 2019; Qiu et al., 2020) Probably, this COVID19 adds to the pre-existing stressors.(Bhatia, 2019; Ivanoff et al., 2018) The significance of gender, was lost in multivariate analysis except for PSS High to low stress comparison, where female endodontists are twice likely to experience high stress than their male counterparts. However, in CPDI, there is no significance.[Table-3]

Age group had a statistically significant association with risk of PS/distress, in both the PSS and CDPI. It was found that younger age groups <25 years, 25-35 years had 3 to 5 times higher risk of moderate and severe PS (PSS scale) in comparison with older age group (>45 years). Interestingly, 25-35 years age group experienced relatively more stress than <25 years group. Similarly, according to CPDI, comparatively to >45 years group, <25 years and 25-35 years had 9 and 4 times higher risk of severe distress respectively. This is not in concordance with Chinese public where, older population had higher distress.(Qiu et al., 2020) It has been reported that age is an important factor in stress and stress reaction.(Knipe et al., 2018; Lo Sasso et al., 2015; Mahal and Shah, 2006) In India, as the age advances, endodontists, possibly acquire financial security and thus have limited stress reaction as opposed to their younger counterparts, who are in the process of financially establishing themselves. The financial uncertainty that the present COVID19 brings could be a possible reason to this phenomenon.(Farooq and Ali, 2020)

Area of practice was significant in association for PSS while for CPDI it was not. [Table-2] India is known to have skewed distribution of dentist and the urban–rural divide exists for a variety of reasons. (Mahal and Shah, 2006) Possibly, the variation in demographics and dominant stressors could also contribute to this phenomenon. As there was not much significance, this factor was eliminated from further analysis.

Experience post-MDS and age appears to be highly related in terms of clinical decision making skills and job satisfaction, perhaps that comes with experiencing different clinical situations and facing odds - clinically as well at personal front. Hence, with increasing experience, endodontists would be more likely to handle stress efficiently than a novice. (Alani et al., 2011; Dechouniotis et al., 2010) In the present study, experience in endodontics had significant magnitude of association, especially postgraduate students and <5 years of experience. Since experience post-MDS and age were collinear, this was not included in subsequent model analysis.

Practice setting had a significant influence on the model wherein Endodontists who were exclusively consultants had constantly expressed increased risk for perceived stress (RRR=5.15 to 8.75, in various situations) as compared to their peers. Practice setting plays a vital role in stress. The stress and job satisfaction levels in group practice or those with multiple models of endodontic practice as compared with solo practice to consultants may vary.(Lo Sasso et al., 2015) As compared to stand alone practice endodontists, those pursuing exclusively consultation (more than 1 clinic) had 3 times elevated risk of severe PS/distress levels in both of PSS or CDPI. This probably indicates that financial implication posed by COVID19 situation is a driver of the (dis)stress. This is further highlighted by the significant difference in PSS and CDPI scores during phase-1 and after announcement of phase-2, when the financial ramification increased with extension of lock down.

All other practice settings, excluding postgraduate students (trainees) and consultants, would have additional avenues of income. Endodontists who rely exclusively on consultations, would not have means or would find it very difficult to practice endodontics during COVID19 situation. This sudden, downward spiral in income could be major driving force for the elevated (dis)stress in this subgroup. It is also observed that trainee endodontists also have elevated risk of stress as compared to solo practitioners. It has been documented that the dental students have higher stress.(Knipe et al., 2018) In the present situation, shifting to a different platform of education, risk of contagion, fear or uncertainty of education and future practice avenues may cause an acute stress on the endodontic trainees/residents. This is captured by the PSS while in CPDI, they have a RRR of 1.67 (mild-moderate to normal) and 0.35 (severe to normal) as compared to solo practitioner, albeit without statistical significance, indicating this stress may not be exclusive to COVID19.(Knipe et al., 2018) In the model age, gender, and practice setting had varying degree of risk with statistical significance.

In COVID19 situation, as compared to Indian public(34%), Indian endodontists in present cohort exhibit higher degree of perceived stress and about 4 in 5 Indian endodontists are perceiving COVID19 to be a major stressor.(Roy et al., 2020) The CPDI model analysis indicate the age and practice setting are major determinants of the distress. It could be postulated that the financial insecurity and fear for getting COVID19 infection through dental operatory aerosol, as one of the possible major driving force of the distress among endodontists.

There is also news report of brewing concerns among Indian public for the need of dental clinics to carter for their urgent oral care needs. However, such reports even highlight the risk of COVID19 infections being spread through dental operatories. (https://www.hindustantimes.com/india-news/covid-19-impacts-dental-surgeries-patients-forced-to-suffer-in-pain/story-O7C5iMFVYNG9SOITTQ4teK.html.Last accessed on 20 April 2020) It has been documented that the Indian dentists had been exposed to COVID19 patients, even without informing authorities as mandated. (https://www.newindianexpress.com/states/telangana/2020/apr/05/dental-hospital-in-ranga-reddy-shut-for-treating-coronavirus-patient-without-informing-officials-2125985.html Last accessed on 20 April 2020) Such dental establishments were legally closed. There are reports in news media of COVID 19 positive cases being treated by dentist or at dental hospital premises. https://www.indiatvnews.com/news/india/hyderabad-woman-found-positive-for-coronavirus-after-death-604551 Last accessed on 20 April 2020) Given this scenario, these news could incrementally add to the existing cynic concern among Indian endodontists. Even otherwise, stress is common in Indian dentists.(Sebastian et al., 2018) This distress (stress and distress often used interchangeably, in present context), could be a transient phenomenon related to COVID19. This may subside when the COVID19 situation dampens or when endodontists learn and adapts to COVID19. But if this distress is in excess, as observed in about 13% of endodontists or persists well after COVID19 fades, it would need to be viewed as a part of a pathological psychological process, which would need professional help.(Phillips, 2009) The impact of the COVID19 on Indian endodontists with features of insomnia, irritability, anxiety, depression, somatoform manifestation has to be carefully screened to identify and treat evolving mental disorders, especially in the most vulnerable ones.

The limitation of this study would include self-reporting bias, non-consideration of Indian states (as the whole country was under lockdown, unlike China), role of other stressors and COVID19 exposure. Further studies, in this direction have to explore role of other possible confounders as well as factor that could meaningfully add to present models.

## CONCLUSION

Psychological (dis) stress among Indian endodontists in COVID19 situation are presented for the first time, to the best of our knowledge. As compared to public, there is 235% more prevalence of stress among Indian endodontists. Those endodontists that are either trainee or those who exclusively go for consultations, females and younger aged have higher risk for (dis)stress. The endodontic and dental key opinion leaders need to factor in the high degree of (dis)stress among endodontists, evolve policies and guidelines such that there is provision for care after the initial coping. They need to work with authorities to develop and coordinate profession focused psychological first aid with adequate space for monitoring, screening, referral and targeted intervention such that the (dis)stress is reduced. The focus should be developing COVID19 lock down exit strategies for dentists, formulating better working protocols for sterilization/disinfection, cheaper, faster, reliable screening and diagnostic tools, making barrier materials available and a robust mental health support system.

## Data Availability

If called upon, the raw data can be shared with consent of all authors

